# Beta-Validation of a Non-Invasive Method for Simultaneous Detection of Early-Stage Female-Specific Cancers

**DOI:** 10.1101/2023.10.06.23296638

**Authors:** Sudhakar Ramamoorthy, Saranya Sundaramoorthy, Ankur Gupta, Zaved Siddiqui, Ganga Sagar, Kanury V.S. Rao, Najmuddin Saquib, Surapeni Krishna Mohan

## Abstract

**Objectives:** In this report, we assessed the accuracy of our previously developed method for simultaneous diagnosis of the four female cancers of the Breast, Endometrium, Cervix, and Ovary, in a clinical set-up with blinded protocol.

**Materials and Methods:** Our test protocol combined global serum metabolome profiling wherein data was analyzed with machine learning algorithms to extract metabolite signatures that correlated with early-stage cancers. High-resolution mass spectrometry was employed to profile the serum metabolome and the resulting data were subjected to a pre-processing pipeline to obtain the data set. The data was then analyzed using artificial intelligence algorithms to identify early-stage cancer metabolic signatures.

**Results:** Overall, a total of 1000 blinded samples were analyzed by generating the serum metabolome profiles, followed by sequential algorithms for cancer detection and multiclass cancer type identification. Of these 1000 samples, 797 were identified as cancer positive, while, 203 samples were identified as cancer-negative. The multiclass algorithm was then applied to the 797 cancer-positive samples, to distinguish between samples that were from patients with either endometrial, breast, cervical, or ovarian cancer. After completion of the analysis, the sample code was broken to estimate the accuracy of the results. Concerning the identification of samples that were cancer-positive, the sensitivity obtained was 99.6% whereas the specificity was 100%. For the second stage of analysis which involves ‘tissue of origin’, all 107 breast cancer samples were correctly identified without any false calls. The accuracy for identification of cervical and ovarian cancers was between 95-96% for each, whereas 91% for endometrial cancer.

**Conclusions:** Our present study validates the performance of our method for the early-stage detection of female-specific cancers in a clinical setting. Importantly, the algorithms for cancer detection and ‘tissue of origin’ prediction, which were initially trained using samples from Caucasian patients, retained the accuracy on samples from Indian women patients. This suggests that the performance of these algorithms was minimally influenced by variables such as ethnicity and race. Present results, therefore, also underscore the potential clinical utility of our method for early-stage diagnosis of cancers that are specific to females.

## INTRODUCTION

Worldwide over 6 million new cancer cases and close to 4 million cancer related deaths are reported every year (1,2). The most prominent female-specific cancers are those of the breast, cervix, endometrium, and ovary (1,3). Breast cancer is the most common and represent ¼ th of cancers in women. (4), whereas cancer of the endometrium and ovary account for 5% and 4% respectively (5,6). Cervical cancer ranks fourth with an estimated 500,000 cases globally (7).

These cancers are often detected at their more advanced stages of progression. Consequently, prognosis is usually poor (2,8,9). While treatment outcomes are significantly improved when the cancer is detected at an early stage (10), effective screening paradigms for early-stage female-specific cancers are either not currently available or suffer from inherent limitations that restrict their scope (11, 12). For example, while mammography is the recommended method for breast cancer screening, it is not a perfect test because its overall sensitivity ranges only from 75% to 85% (13). The sensitivity further decreases sharply in the case of dense breast parenchyma (14). The net consequence of these limitations is that many women with breast cancer are missed (14). Another downside to mammography is its relatively high false-positivity rate, which is further enhanced in women below the age of 40 years. As a result, the likelihood that a woman will receive at least 1 false-positive call after 10 yearly mammograms is found to be as high as 61% (15). It follows, therefore, that an improved breast cancer screening method with high sensitivity and specificity across all age groups of women is clearly needed to minimize the harm to benefit ratio and, thereby, obtain better outcomes. Efforts to achieve this are presently ongoing (16)

Current guidelines recommend three primary screening options for cervical cancer. These are cytological testing alone, standalone high-risk human papilloma virus (hrHPV) testing, and co-testing with the combination of cytological and hrHPV testing (17). While cytological testing is more widely accepted as the primary screening method for cervical cancer (18), it suffers from poor sensitivity (47-70%) although the specificity is high (19). In contrast, hrHPV standalone testing yields higher sensitivity (80%), but with a higher false positivity rate of about 15% (20,21). While sensitivity can be improved upon co-testing with both methods this, however, was shown to occur at the cost of lower specificities (20). In contrast to at least some screening methods being available for breast and cervical cancer, no such routine or standard screening test is currently available for either endometrial or ovarian cancer. Indeed, most ovarian cancer patients do not experience symptoms until its later stages. The only recourse currently available is to either measure levels of the relatively non-specific tumor marker CA 125 in the blood, or to perform a pelvic ultrasound. However, because of poor sensitivity especially for early-stage cancer, and a high false-positivity rate, neither of these tests currently qualify as standalone screening tests for ovarian cancer (22,23). These collective results clearly highlight the need for developing more accurate and reliable methods for screening of each of these cancers. The ideal solution here would be a non-invasive test that would enable more effective surveillance of at least the more vulnerable segments of the female population.

In an earlier report we had described the development of a method for the concomitant early-stage detection of these female-specific cancers with very high accuracy. Our protocol combined untargeted metabolomics with machine learning based data analysis to extract metabolite patterns that define the early-stage cancers (24). The accuracy of detection obtained across all four cancers was 98% while a subsequent analysis also helped to identify the cancer type of the individual cancer-positive samples with reasonable accuracy. For the development of this test, we had employed stored serum samples that were obtained from biobanks. To evaluate its potential for clinical use, however, it was important for us to ascertain its performance with samples that were directly obtained from women patients in a clinical setting. The present report describes our efforts in this direction. Results obtained establish that the fidelity of both cancer detection and prediction of the tissue of origin (TOO) remained exceptionally high upon analysis of serum samples collected directly from women volunteers. Importantly, we were also able to obtain beta-validation of our method by confirming its accuracy in a blinded study protocol.

## MATERIALS AND METHODS

### Study design and sample collection

Blood, from both cancer patients and normal female volunteers, were collected at the Panimalar Medical College Hospital and Research Institute after first obtaining approval from the Panimalar Medical College Hospital & Research institute - Institutional Human Ethics Committee (Protocol No. PMCHRI-IHEC-034). Serum was subsequently prepared, and the samples were anonymized before submitting to the analytical team (AG, GS, ZS, KVSR, NS) for analysis.

### Sample Accessioning

Unique identity number (identifier) issued to each sample post arrival to lab. These identifiers were used for queries related to sample handling, tasks, and results. Samples were stored frozen (-80°C) until required.

### Metabolite extraction

Extraction of metabolite was done as earlier reported by Gupta *et*.*al* (24). Briefly, serum samples were thawed at 4C on ice. 10µl of each sample was taken and metabolites were extracted and processed for UHPLC-MS/MS as previously described (24).

### Liquid Chromatography coupled with high resolution Mass Spectroscopy

The detailed method of untargeted metabolomics was previously reported by Gupta *et*.*al* (24). Briefly, a Dionex LC system was employed for the high-resolution separation of serum metabolite in normal and cancer patients.

10ul of each sample was resolved by UHPLC prior to MS/MS analysis as previously described (24).

### Data processing

Data generated by mass spectrometers was pre-processed through the steps of mass error correction, ion filtering, and normalisation as described earlier by Gupta *et*.*al* (24).

#### AI modeling of the data

Detailed explanation of the method was shown by Ankur *et*.*al* (24). Briefly, a layered approach was used, where we first distinguish cancer from normal and then between each of the cancers. The scheme employed for training, testing and validation along with method used for determining sensitivity and specificity was as previously described (24).

## RESULTS

### Study rationale and the samples employed

In our previous study (24) we had developed a method, by integrating untargeted serum metabolomics by mass spectrometry and analysis of the resulting data with a machine learning algorithm to diagnose Stage 0/I of female cancers with high fidelity. These were breast, endometrial, cervical and ovarian cancers. Our protocol involved a sequential approach in which the cancer-positive samples were first distinguished from the non-cancer samples. Subsequent to this, multi-class algorithm was employed to differentiate between each of the separate cancers of the cancer-positive subset. The high accuracy obtained in this study suggested the possibility that this approach could indeed be further developed as a screening tool for the early-stage female-specific cancers.

Although our results were extremely promising, we recognized that our analysis algorithms had been developed using serum samples obtained from commercial biobanks (24). Therefore, to further explore its utility, it was necessary to evaluate the performance of our test under more relevant conditions where samples were directly obtained from patients in a clinical setting. Another important point here was that, for the algorithm development, we had employed metabolome profiles of sera that had been primarily derived from Caucasian patients. Therefore, since the metabolome composition is known to vary across different populations (28, 29), it was also relevant to establish whether these algorithms were equally effective when samples obtained from Indian women patients were tested. This question was especially pertinent given that our objective was to identify metabolite signatures that specifically characterized the disease state of interest, independent of other variables such as age, ethnicity, race, etc. We, therefore, undertook the present exercise to ascertain the accuracy of our method (24) when tested on samples directly obtained from Indian women patients.

### Verification of algorithm performance for identification of Indian women patients

Our first objective was to test the performance accuracy of our earlier developed algorithms, which were developed using data largely generated from Caucasian samples, in Indian patients. For this we used a set of 300 samples collected at the Panimalar Medical College Hospital and Research Institute (PMCHRI). Of these, 71 samples each were from patients with endometrial, cervical, or ovarian cancer, where the remaining 87 samples were from breast cancer patients. In addition, we also included 200 samples from normal volunteers. The distribution across age groups, BMI status, and cancer stages are given in Table-1.

**Table 1:** Demographic, BMI, and Cancer-stage grouping of the samples employed for algorithm verification.

| Sample Clinical Info |  |  |  |  |  |  |
| --- | --- | --- | --- | --- | --- | --- |
| Parameter | Intervals | Clinical Status |  |  |  |  |
|  |  | Normal control<br>(n=200) | Endometrial cancer<br>(n=71) | Breast cancer<br>(n=87) | Cervical cancer<br>(n=71) | Ovarian cancer<br>(n=71) |
| Age(years) | 20-30 | 102 | 0 | 8 | 1 | 5 |
|  | 31-40 | 51 | 2 | 26 | 5 | 17 |
|  | 41-50 | 29 | 17 | 19 | 7 | 27 |
|  | 51-60 | 14 | 35 | 28 | 24 | 16 |
|  | 61-70 | 4 | 13 | 3 | 20 | 5 |
|  | 71-80 | 0 | 4 | 3 | 13 | 0 |
|  | 81-90 | 0 | 0 | 0 | 1 | 1 |
| BMI (kg/m2) | 10 to 30 | 154 | 57 | 68 | 57 | 56 |
|  | > 30 | 24 | 14 | 19 | 12 | 15 |
| Cancer stage | I | - | 26 | 32 | 29 | 21 |
|  | II | - | 38 | 46 | 34 | 32 |
|  | III | - | 4 | 3 | 3 | 11 |
|  | IV | - | 3 | 6 | 5 | 7 |

Metabolites were extracted from each of the samples and then analyzed as previously described (24). Range of the unique metabolite numbers detected in normal control samples and the individual cancers, across the individual age groups, is depicted in Figure 1. The data was subsequently processed using our previously described in-house pipeline to eventually extract the matrix consisting of 2764 features (24).

**Figure 1.**
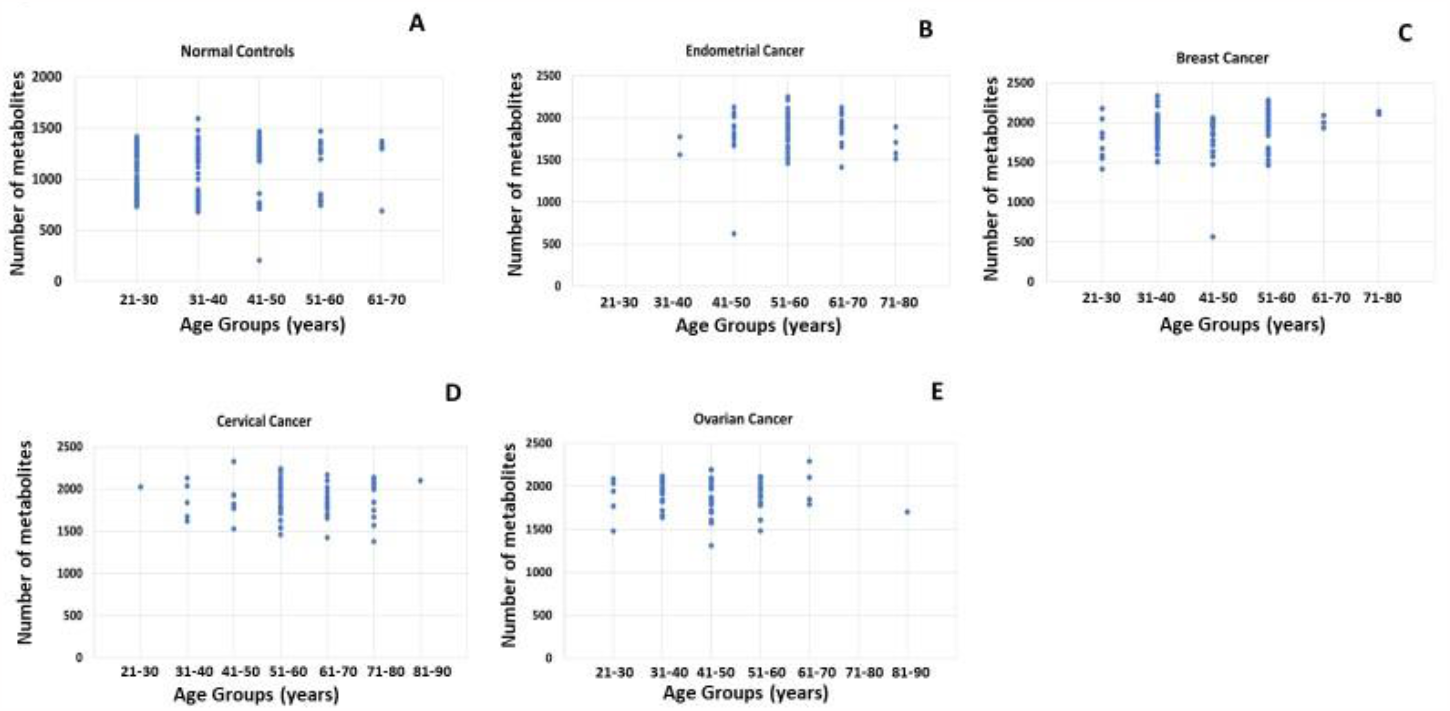
Age-wise detection of detected metabolites. Figure provides a graphical representation of the number of metabolites detected across the individual age groups, for the normal control set as well as the individual cancer groups. The cumulative unique metabolites detected in normal control samples were 5371. While, endometrial, breast, cervical and ovarian cancer samples were found to have 4947, 5132, 5035 and 5041 respectively.

The PLSDA plot generated using the matrix is shown in Figure 2. It is evident from this figure that the cancer samples could be unambiguously separated from the normal controls. To differentiate the cancer samples from normal controls, we applied our previously developed AI model (24) on the present data as the test set. As described in our earlier report (24), the model applies a logistic regression function to separate the female-specific cancer samples from the control group. After training the algorithm calculates a score for the individual samples by using the following formula:

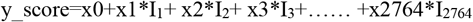

**Figure 2.**
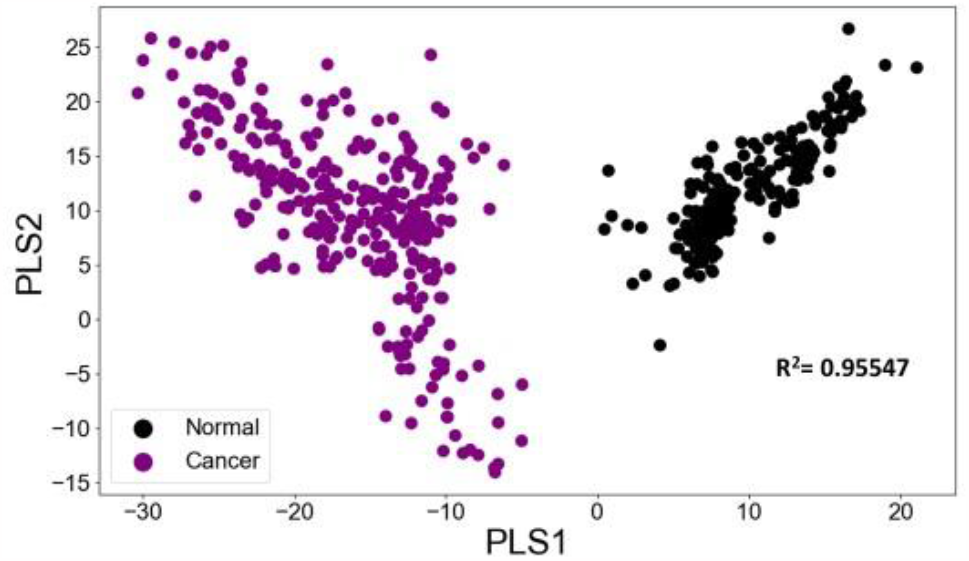
PLSDA plot distinguishes between the cancer group and also the normal controls. Figure presents a PLSDA plot of the matrix of sample-specific metabolites versus metabolite intensity for normal controls and the group of women-specific cancer samples. The separation obtained between them is shown. The R^2^ value obtained is included.

Here, x0 is a constant number, I_i_ (1<=i<=2764) is the intensity of metabolite i present in the respective sample.

The schematic of the approach employed is illustrated in Figure 3 and Figure 4A shows the results obtained after applying the model on the test set (24). The scatter diagram gives the scores generated by the model for control and the cancer cases. It is evident from the figure that the scores for the normal control samples are clearly distinguished from those for the cancer set. A cut-off score of 5 successfully differentiated the two sets as depicted in Figure 4B. Sensitivity, Specificity, and Accuracy all corresponded to 100%.

**Figure 3.**
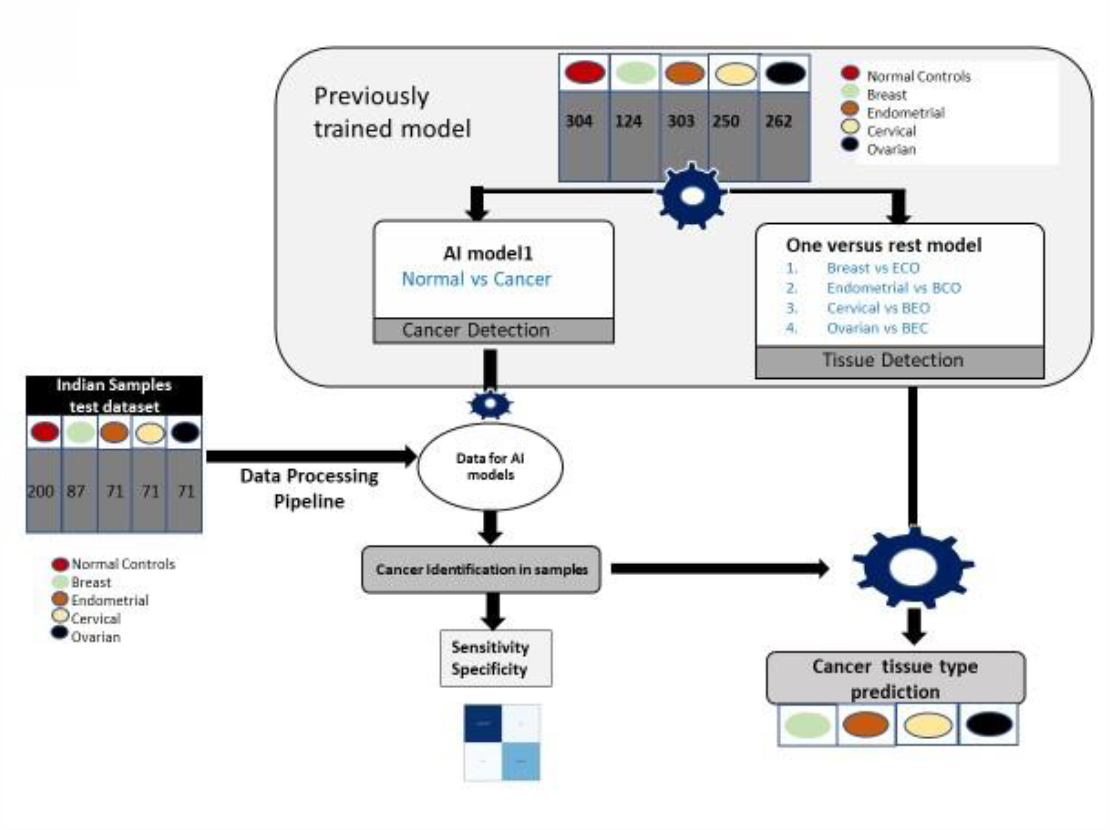
Schematic of the approach employed for identifying women-specific cancer positive samples in the Indian test dataset. Illustrated here is the procedure adopted for identifying the cancer-positive samples in the Indian Test dataset. The grey box represents the previously trained model for cancer detection and tissue type identification (24). The Test dataset was first processed using the data processing steps followed previously in the study (24). The final processed data was tested first for the identification of cancer samples in the dataset, the cancer positive identified cases were next further tested using the one versus rest models trained previously for cancer tissue type prediction (24).

**Figure 4.**
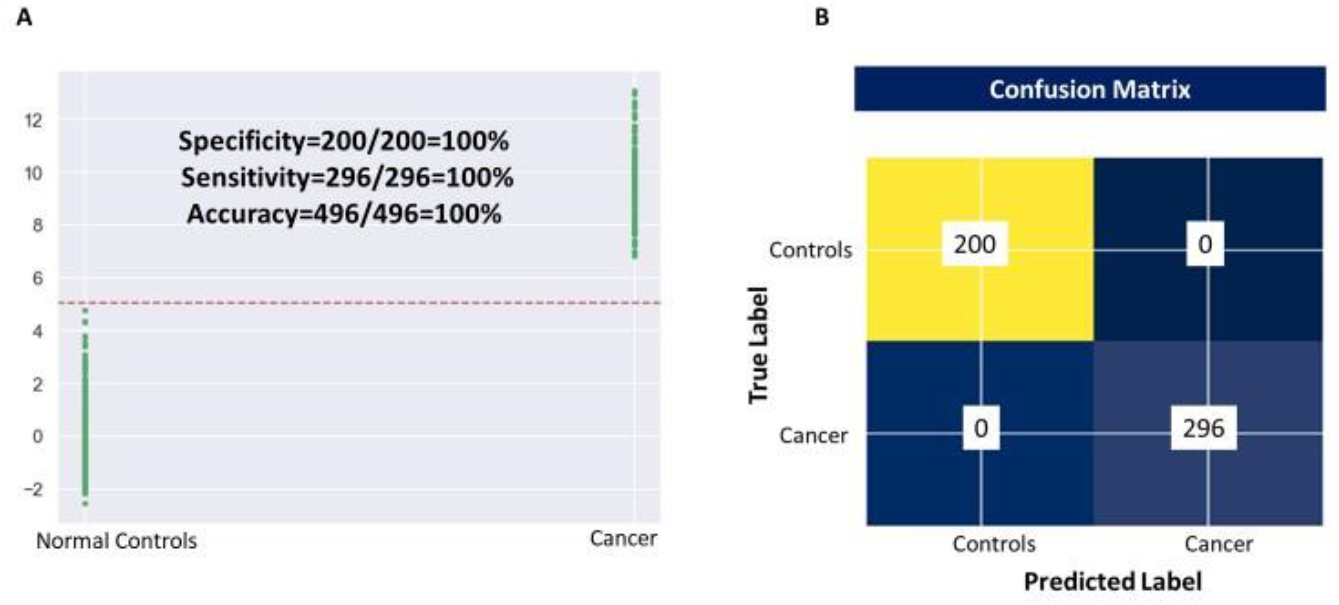
Distinguishing women-specific cancers from normal controls. Results obtained for testing of our previous trained model for distinguishing the women-specific cancer group from normal controls (24) on the data set generated from Indian patient and normal controls is shown here. The separation achieved between the cancer and normal control groups is shown in Panel A and the resulting confusion matrix that was generated is shown in Panel B. Values obtained for Sensitivity, Specificity, and Accuracy are also given.

### Differentiating between Breast, Endometrial, Cervical and Ovarian cancer samples

Next, to distinguish the specific cancer types among the positive samples identified in Figure 4, we layered the multiclass AI model over the model described in this figure. The multiclass AI model was also developed in our earlier study (24). It is a one versus rest (OVR) classifier model, which gives four scores for each sample. Each of these scores define the likelihood of a given sample belonging to anyone of the four classes. As previously described (24), the formulae used by the algorithm to calculate the four scores for each of the sample is as follows:

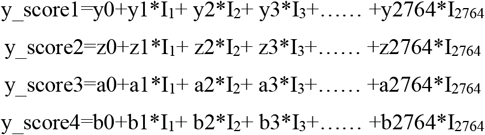

y0, z0, a0, b0 represent constant numbers, I_i_ (1<=i<=2764) is the relative concentration of metabolite i in the respective sample.

To assess the performance of our multiclass model in terms of differentiating between the individual cancer types for Indian patients, as also from normal controls, the scores obtained from the model were plotted. In Figure 5A, we plotted the score obtained for Endometrial cancer samples against that of the group comprising of the Breast, Cervical and Ovarian (BCO) cancers. A clear distinction between Endometrial and BCO Cancer samples is seen (Fig. 5A). Control samples were also included in this analysis and sensitivity, specificity and accuracy obtained were 97%, 94%, and 95% respectively.

**Figure 5.**
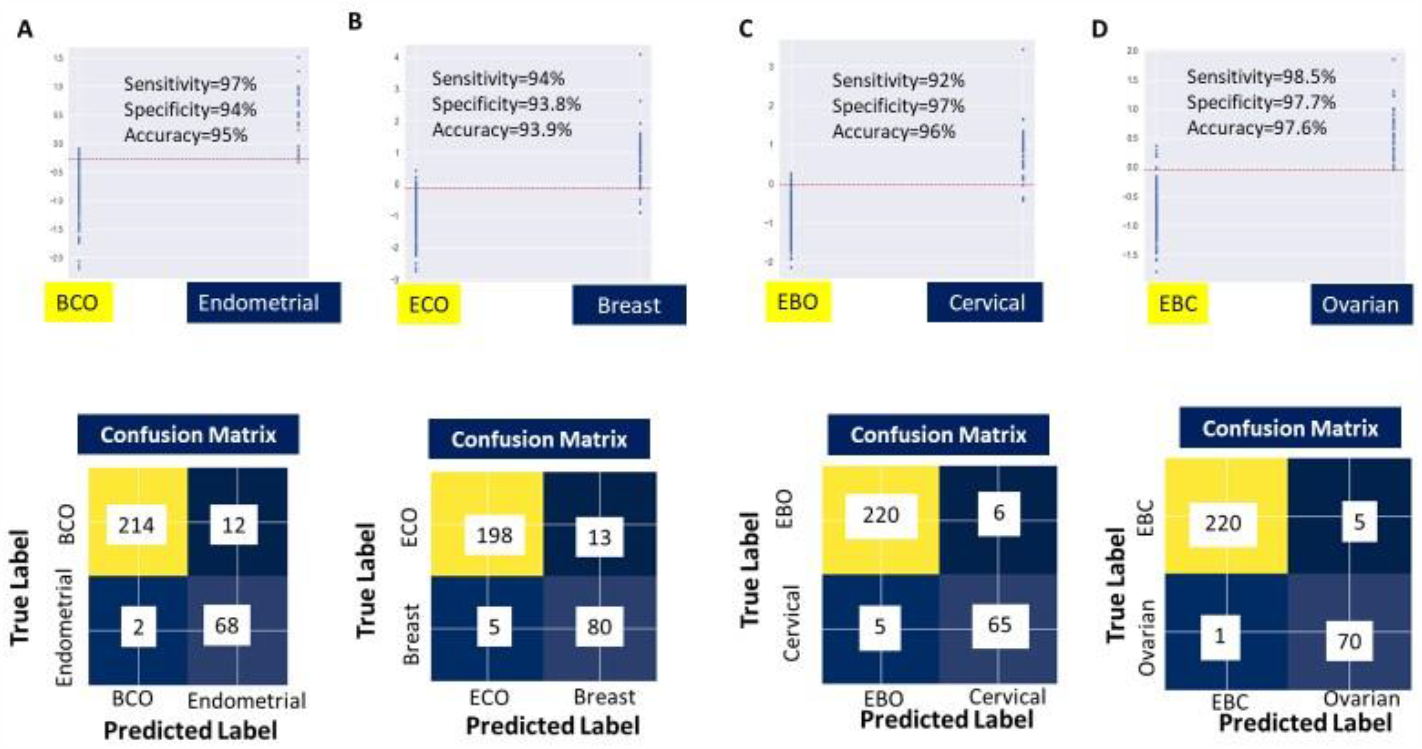
Testing the multiclass model for its ability to distinguish the individual cancer groups. Panel (A) shows the results of specifically testing the multiclass trained model for separation of endometrial cancer samples from the other cancers (breast, cervical, ovarian; abbreviated as BCO) based on model’s Endometrial scores. The resulting confusion matrix on applying a threshold shows good accuracy, sensitivity, and specificity. Panel (B) shows the results of specifically testing the multiclass trained model for separation of breast cancer samples from the other cancers (endometrial, cervical, and ovarian; abbreviated as ECO) based on model’s Breast scores. The resulting confusion matrix on applying a threshold shows good accuracy, sensitivity, and specificity. Panel (C) the results of specifically testing the multiclass trained model for separation of cervical cancer samples from the other cancers (breast, endometrial, ovarian; abbreviated as EBO) based on model’s Cervical scores. The resulting confusion matrix on applying a threshold shows good accuracy, sensitivity, and specificity. Panel (D) shows the results of specifically testing the multiclass trained model for separation of ovarian cancer samples from the other cancers (breast, endometrial, cervical; abbreviated as EBC) based on model’s Ovarian scores. The resulting confusion matrix on applying a threshold shows high accuracy, sensitivity, and specificity.

Next, we plotted the Breast cancer sample scores versus those of the group comprised of Endometrial, Cervical and Ovarian (ECO) cancer samples (Fig. 5B). Here again Breast Cancer could be clearly distinguished from ECO cancer sample set as shown (Fig. 5B). The sensitivity, specificity and accuracy obtained were 94%, 93.8%, and 93.9% respectively.

Figure 5C depicts the confusion matrix obtained for an analysis of Cervical cancer sample scores versus that for the group of Endometrial, Breast and Ovarian (EBO) cancer samples. Sensitivity, specificity and accuracy obtained were 92%, 97%, and 96% respectively. And, finally, Figure 5D shows the confusion matrix obtained for discriminating Ovarian cancer samples from the group comprising of Endometrial, Breast and Cervical (EBC) Cancer samples. Here, the calculated the sensitivity, specificity and accuracy were 98.5%, 97.7%, and 97.6% respectively.

### Beta-validation of our algorithms through a blinded study protocol

The results obtained so far establish that the algorithms that we had previously developed, using data primarily generated with samples obtained from Caucasians (24), were equally effective for screening of female-specific cancers in Indian women. Indeed the high sensitivity and specificity obtained, for both detection of cancer positivity as well as for identification of cancer type, also suggest that this approach is likely to be at least less sensitive to population-related differences in the subjects. To further verify the potential clinical utility of our test platform, we sought to beta-validate it by employing a blinded protocol. In this, coded samples we sent by the clinical team (SR, SS, SKM) to the team involved in sample analysis and diagnosis of the cancer state and type (analytics team, AG, GS, ZS, NS, KVSR). Here, all public health information (PHI) identifiers/variable were protected from the latter team. Apart from sample IDs, no information on either cancer status or cancer type were provided.

A total of 1000 samples were provided by the clinical team, which the analytical team then analysed by generating the corresponding serum metabolome profiles, followed by sequential application of the cancer detection and multiclass cancer type identification algorithms. Of these 1000 samples, 797 were identified as cancer positive, whereas the remaining 203 samples were identified as cancer negative. Following this, the multiclass algorithm was then applied to the 797 samples putatively identified as cancer positive, in order to distinguish those samples that were patients with either endometrial, breast, cervical, or ovarian cancer. The cumulative results were then sent to the clinical team, which then broke the code and summarized their findings. These findings are presented in Tables 2 and 3 where Table 2 provides the results generated with the first algorithm for identification of cancer positive samples. It is evident from this table that a very high detection accuracy was obtained, with no false positives and only 3/800 true positive samples being missed as false negatives. The sensitivity obtained here was 99.6%, with a specificity of 100% (Table 2). Table 3 presents the results obtained after application of the multiclass algorithm for distinguishing between samples from patients with either breast, endometrial, cervical, or ovarian cancer. Here, the algorithm was only applied to the 797 blinded samples that were identified as cancer positive in Table 2. The results in Table 3 clearly underscore the high degree of fidelity with which the individual cancer types could be distinguished. While all 107 of the breast cancer samples were correctly identified, the accuracy for identification of cervical and ovarian cancer samples was between 95 – 96%. Endometrial cancer identification was, however, somewhat lower with an accuracy of close to 91% (Table 3). These results, therefore, provide strong validation for our method, which integrates untargeted metabolome profiling with AI-driven data analytics, for concurrent detection of female-specific cancers.

**Table 2:**
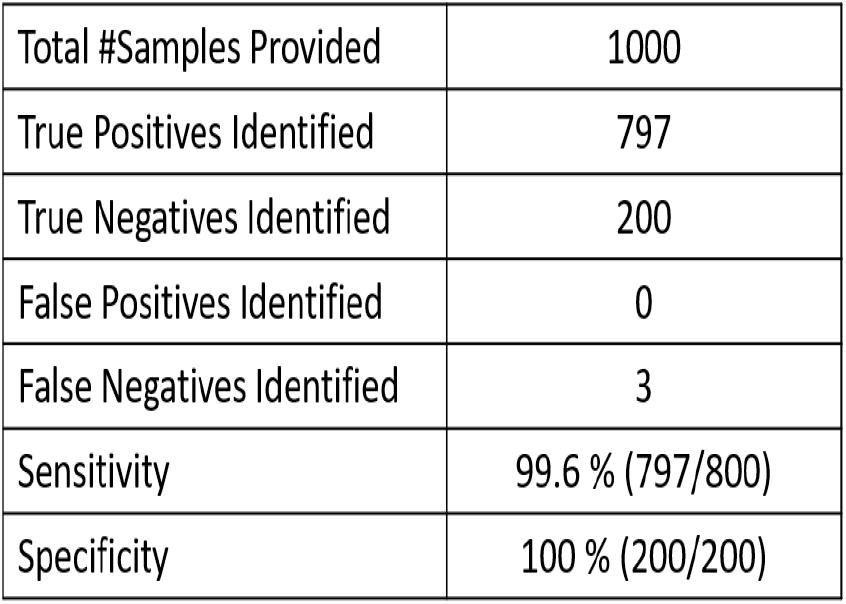
Identification of women-specific cancer patients from the blinded sample set.

**Table 3:** Identification of the cancer type (TOO).

| **Cancer Type** | **#Samples correctly identified** | **Accuracy (%)** |
| --- | --- | --- |
| Breast Cancer | 107/107 | 100 |
| Endometrial Cancer | 272/299 | 90.97 |
| Cervical Cancer | 184/192 | 95.8 |
| Ovarian Cancer | 190/199 | 95.5 |

## DISCUSSION

While there is an increased emphasis on detection of cancers at an early stage, recent years are also witnessing a paradigm shift towards multiple cancer detection with a single blood draw (30). These latter efforts have largely focused on analysis of circulating tumour cell-free DNA (ctDNA) by using genomic technologies and machine learning to not only detect multiple cancers simultaneously but also identify the tissue of signal origin (TOO) (31-35). The promise that such multi-cancer detection (MCD) tests hold is that they would permit detection of several cancers that otherwise go undetected until they reach an advanced stage (36). Indeed, mathematical modelling of the potential public health impact of including multi-cancer detection tests to usual care suggested a marked positive effect with a substantial reduction in overall cancer mortality (37).

Despite the promise shown by MCDs based on ctDNA analysis, concerns have been raised that ctDNA may be a less than satisfactory proxy for liquid biopsies of tumour tissues for early detection because of limitations in both sensitivity and specificity (38, 39). In this context, the low concentration of ctDNA and its variability based on type and status of the tumour also contributes as a complicating factor (40). Furthermore, the low to negligible concentration of ctDNA present in the early stages of cancer also limits the sensitivity that can be achieved for the stages (41, 42). This concern is borne out by more recent results from clinical trials where sensitivity of detection of early-stage cancers was indeed found to be low (34, 35). Given these potential limitations with ctDNA, we preferred to test the utility of metabolomics for early-stage cancer detection. The rationale here was based on the now widely accepted notion that metabolites serve as proximal reporters of disease since their relative abundances are often directly related to pathogenic mechanisms (43-45). We hypothesized that metabolomics should be an especially relevant technique for cancer detection since cancers have significantly altered metabolism (46, 47). Therefore, the spectrum of metabolites produced can possibly yield signatures characteristic of the presence of cancer.

As demonstrated in our previous report (24) and the results described here, our expectation was indeed borne out. At least in the context of the four female-specific cancers, serum metabolome analysis coupled with AI analysis yielded a very accurate method for their simultaneous detection at the early stage, and subsequent of the tissue of origin (TOO). Indeed the high sensitivity obtained for detection of Stage-I of all the four cancers is particularly notable and unmatched by that achieved through analysis of ctDNA (34, 35). We do, however, acknowledge that our current method only enables simultaneous detection of the four female-specific cancers. Nonetheless, an important highlight of our present study is that it validates the performance of our method in a clinical setting. Particularly notable here is the fact that the algorithms for cancer detection and TOO prediction, which were trained primarily on samples from Caucasian patients (24), retained their accuracy with samples from Indian women patients. This indicated that, at least with respect to the two population groups studied, variables such as ethnicity and race did not affect performance of either algorithm. Indeed, the high sensitivity and specificity of >99% obtained for detection of the cancer positive samples, and the high TOO prediction accuracy, in the blinded study especially underscore this, while also serving to validate the method developed by us. We do recognize, however, that a more extensive clinical trial would be required to demonstrate its clinical utility. Furthermore, it will also be of interest to explore whether early detection of additional cancers can be brought within the ambit of this approach.

## Data Availability

All data produced in the present study are available upon reasonable request to the authors.

## AUTHOR CONTRIBUTIONS

S.R, S.S., and S.K.M. coordinated the procurement of patient samples and confirmation of their diagnosis. N.S., G.S., and Z.S. led all experimental aspects of the study while data analysis was performed by A.G. The study was jointly supervised by K.V.S.R. and S.K.M. N.S., K.V.S.R., and S.K.M. wrote the paper.

